# The importance of increasing primary vaccinations against COVID-19 in Europe

**DOI:** 10.1101/2023.01.09.23284297

**Authors:** Pierre-Yves Boëlle, Eugenio Valdano

## Abstract

In the European Union, mass vaccination against COVID-19 staved off the strict restrictions that had characterized early epidemic response. Now, vaccination campaigns are focusing on booster doses, and primary vaccinations have all but halted. Still, 52 million European adults are unvaccinated. We investigated if reaching the still unvaccinated population in future vaccination campaigns would substantially decrease the current burden of COVID-19, which is substantial. We focused on vaccination homophily, whereby those who are unvaccinated are mostly in contact with other unvaccinated, making COVID-19 circulation easier. We quantified vaccination homophily and estimated its impact on COVID-19 circulation.

We used an online survey of 1,055,286 people from 22 European countries during early 2022. We computed vaccination homophily as the association between reported vaccination status and perceived vaccination uptake among one’s own social contacts, using a case-referent design and a hierarchical logistic model. We used this information in an analysis of the COVID-19 reproduction ratio to determine the impact of vaccine homophily in transmission.

Vaccination homophily was present and strong everywhere: the average odds ratio of being vaccinated for a 10-percentage-point increase in coverage among contacts was 1.66 (95% CI=(1.60, 1.72)). Homophily was positively associated with the strictness of COVID-19-related restrictions in 2020 (Pearson=0.49, p-value=0.03). In the countries studied, 12%-to-18% of the reproduction ratio would be attributable to vaccine homophily.

Reducing vaccination homophily may curb the reproduction ratio substantially even to the point of preventing recurrent epidemic waves. In addition to boosting those already vaccinated, increasing primary vaccination should remain a high priority in future vaccination campaigns, to reduce vaccination homophily: this combined strategy may decrease COVID-19 burden.

## Introduction

With mass vaccination, countries in the European Union (EU) are now managing COVID-19 without resorting to the pervasive closures and movement restrictions that characterized the first year and a half of the pandemic. Vaccine uptake in the EU is high: 86% of adults have received at least one dose^1^. There, COVID-19 is transitioning from being an emergent disease to an endemic respiratory disease imposing considerable morbidity and mortality^2^. Vaccines will likely remain the most effective public health tool for its routine management, as treatment options are limited^3,4^, and pervasive non-pharmaceutical interventions are no longer sustainable or socially justifiable. But what should future vaccination campaigns against COVID-19 aim for? Two extreme targets could be envisioned: The first is to increase the number of primary vaccinations among those 52 million adults who never got vaccinated; the second is to boost immunity among those already vaccinated, to make up for the relatively short-lived immunity conferred by vaccines^5^ and to increase their efficacy against new SARS-CoV-2 variants and subvariants. Current campaigns are unquestionably pursuing the latter, as policies to increase primary vaccinations, like vaccine mandates and restrictions selectively targeting the unvaccinated, have been scaled down. Primary vaccinations have stalled since early 2022 and policymakers have little hope, and no plans, to persuade those still unvaccinated to get the jab. Giving up on primary vaccinations, however, may mean accepting the current burden of COVID-19 as part of its long-term management, in terms of incidence of severe disease^6^, mortality - the EU recorded 242,000 COVID-19-attributable deaths from January to October 2022^1^ - and post-acute sequelae^6–8^. In this study we show that increasing the number of primary vaccinations would still be extremely beneficial in terms of epidemic control. This is because in all EU countries under study, those who are not vaccinated tend to be socially connected and cluster together - an effect that may be described as *vaccination homophily*. Vaccination homophily implies that those unvaccinated are in contact with other unvaccinated individuals, even if the overall vaccination coverage is high. This creates clusters with low vaccination uptake and favors more frequent and larger outbreaks, making population-level epidemic control harder. This phenomenon has already been reported for flu^9^, measles^10–13^ and pertussis^12,14^ and explored in modeling studies^15–18^. Here, we quantified vaccination homophily for COVID-19 and found that it contributes to a sizeable share of the reproduction ratio - the average number of secondary cases that a case generates. Also, we found that this share increases as the already vaccinated population is boosted. Our study implies that vaccination coverage as a metric of population protection may be misleading and that increasing primary vaccinations – with the consequence of reducing homophily – would have an effect beyond direct protection: It would help bring down incidence in the community, reducing the burden of COVID-19 below current levels.

## Methods

### Survey data and complementary data sources

Data on vaccination was obtained from *The University of Maryland Social Data Science Center Global COVID-19 Trends and Impact Survey*^19^. This survey collects questionnaires filled by Facebook users. We included in the study 21 out of 27 member countries of the European Union, excluding Cyprus, Estonia, Latvia, Lithuania, Luxembourg, Malta, for insufficient data. We also included Norway, a member of the European Economic Area that participates to the network of the European Centre for Disease Prevention and Control (ECDC). Questionnaires were offered in different languages, to Facebook users aged 18 years or older. We included questionnaires from January 1^st^, 2022, to April 30^th^, 2022. In this period vaccination coverage was stable in all countries (see appendix figure S1): the largest increase was in Germany where an additional 2.3% of the adult population received a first injection. Responders were solicited at most once per month. From each questionnaire, we obtained age (recoded as 18 to 34 years old, 35 to 64 years old, 65+ years old), gender and the two following questions: 1) “Have you had a COVID-19 vaccination?”, possible answers: “yes”, “no”; 2) “Thinking about your friends and family, how many have gotten a COVID-19 vaccine?”, possible answers: “None of the people”, “A few people”, “Some people”, “Most people”, “All of the people”. We excluded users who did not reply to either question. Tables 1, 2 report the number of respondents in the study, by country, vaccination status, gender and age. Additional data to complement the analysis were country-level vaccination coverage and COVID-19-related deaths from ECDC^1^ and the Stringency Index from the COVID-19 Government Response Tracker^20^. For the latter, we averaged the daily reported values in each country over the period March 2020 – December 2020, to compute the tightness of restrictions during the 1^st^ year of the pandemic.

**Tab. 1:** Included survey respondents, by country and vaccination status. Percentages are computed on the total in each country, and the overall total in the last row.

| country | non vaccinated | vaccinated | total |
| --- | --- | --- | --- |
| AUT | 4,290 (17%) | 20,573 (83%) | 24,863 |
| BEL | 2,393 (10%) | 22,215 (90%) | 24,608 |
| BGR | 5,608 (37%) | 9,493 (63%) | 15,101 |
| CZE | 4,488 (17%) | 21,391 (83%) | 25,879 |
| DEU | 16,353 (11%) | 133,804 (89%) | 150,157 |
| DNK | 1,510 (4%) | 33,403 (96%) | 34,913 |
| ESP | 3,467 (5%) | 60,309 (95%) | 63,776 |
| FIN | 1,344 (7%) | 16,943 (93%) | 18,287 |
| FRA | 16,126 (12%) | 120,914 (88%) | 137,040 |
| GRC | 3,201 (11%) | 25,857 (89%) | 29,058 |
| HRV | 2,835 (26%) | 7,883 (74%) | 10,718 |
| HUN | 6,306 (14%) | 38,170 (86%) | 44,476 |
| IRL | 821 (6%) | 11,983 (94%) | 12,804 |
| ITA | 8,616 (6%) | 139,345 (94%) | 147,961 |
| NLD | 4,911 (11%) | 38,313 (89%) | 43,224 |
| NOR | 1,691 (5%) | 31,799 (95%) | 33,490 |
| POL | 7,484 (17%) | 36,459 (83%) | 43,943 |
| PRT | 1,981 (5%) | 39,359 (95%) | 41,340 |
| ROU | 7,079 (21%) | 26,548 (79%) | 33,627 |
| SVK | 2,930 (21%) | 10,746 (79%) | 13,676 |
| SVN | 1,415 (23%) | 4,831 (77%) | 6,246 |
| SWE | 5,504 (5%) | 94,595 (95%) | 100,099 |
| **TOTAL** | **110,353 (10%)** | **944,933 (90%)** | **1,055,286** |

**Tab. 2:**
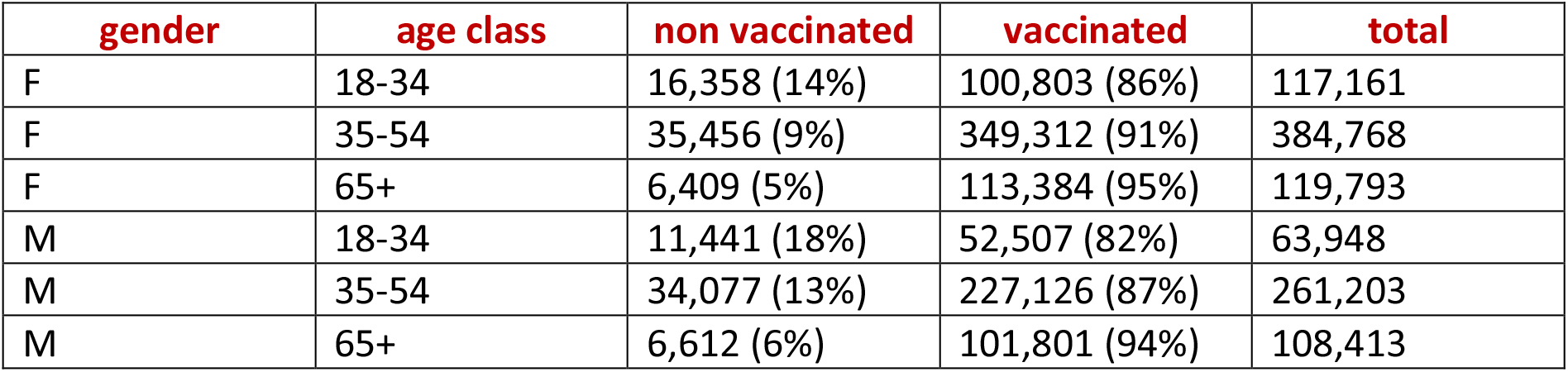
Included survey respondents, by gender, age and vaccination status. Percentages are computed on the total of each row.

### Inferring vaccination homophily

We defined *perceived coverage* for an individual as the fraction of those vaccinated among their friends and family. To measure vaccination homophily, we used a case-referent design, in which cases were vaccinated responders and referents were the unvaccinated, and the exposure was measured by perceived coverage. We assumed a logistic dependency between perceived coverage *η* ∈ [0,1] and vaccination status *v* ∈ {0,1} (with *v* = 1meaning vaccinated) as follows:

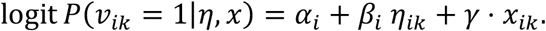

The indices indicate the country (*i)* and the *k-th* the individual of country *i*. Coefficient *β*_*i*_ is the log odds-ratio of perceived coverage for vaccination, i.e., exp(*β*_*i*_Δ*η*) is the odds-ratio of being vaccinated for a Δ*η* increase in perceived coverage. In the following, we will report odds ratios for an increase of 10 percentage points in perceived coverage (Δ*η* = 0.1) and call it *vaccination odds ratio*. The variable *x*_*ik*_ encodes the demographic characteristics of individual *k*, namely gender (Female vs. Male) and age in 3 classes (< 35, 35-64, >= 65) with *γ* the corresponding coefficients. We adopted a hierarchical description for *α* _*i*_, *β*_*i*_as:

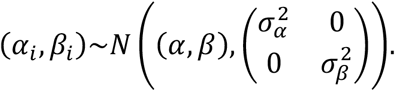

In the data, perceived coverage is available through the ordinal variable *J*={“None of the people”, “A few people”, “Some people”, “Most people”, “All of the people”}. We modeled this as

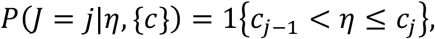

where 1{.} is the indicator function and {*c*} is a set of 4 cutpoints to be estimated defining the boundaries for discretisation of *η*: *c*_0_ = 0 < *c*_1_ < *c*_2_ < *c*_3_ < *c*_4_ < *c*_5_ = 1. This finally gives the individual-level likelihood as follows:

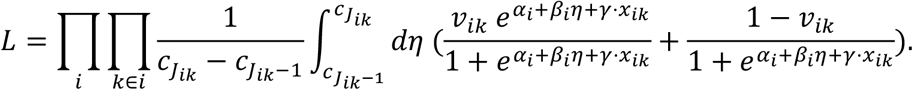

The likelihood is explored using a Markov-Chain-Monte-Carlo (MCMC), with non-informative priors on *α, β, γ*, 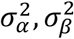 and {*c*}, implemented in Stan 2.1 interfaced with Python 3.8. More on the statistical inference is available in the appendix. Model diagnostics (identifiability) and MCMC diagnostics are available in appendix figure S2.

### Computing correlations with vaccination homophily

We computed the Pearson correlation of vaccination homophily in each country with some other quantities, e.g., the vaccination coverage reported by the ECDC. To account for uncertainty in homophily (coming from the uncertainty in *β*), we computed the Pearson correlation coefficients with each sample of the posterior distribution of the *β*s generated by the MCMC sampling. The median value of these coefficient was then reported as the measure of correlation. To compute P-values, we repeated this approach with permutation of the countries (3000 permutations). The P-value was then computed as the percentage of (permuted) correlation coefficients more extreme than the original measure. Statistical significance was set at the 0.05 threshold for P-values.

### Proportion of the reproduction ratio attributable to vaccination homophily

We computed the reproduction ratio for COVID-19 using the next-generation matrix^21^ structured by age, gender and vaccine homophily. The next generation matrix holds the number of secondary cases that will be directly infected by a primary case in the population, accounting for the structure of the population. Here, we indexed the matrix according to vaccination status and age/gender characteristics (*v, x*) in the primary case and those in secondary cases (*v*^′^, *x*^′^) as *M*(*v, x*; *v*^′^, *x*^′^). The largest eigenvalue of this matrix yields the reproduction ratio *R* (Ref. ^21^). We wrote *M* as the sum of cases arising from contacts obeying vaccine homophily and those mixing randomly as *M* = *M*_*homo*_ + *M*_*rand*_. Each matrix could in turn be split as arising from contacts in households and in the community, informed by age-, gender-stratified mixing matrices of household contacts (*C*_=_(*x, x*′)) and community contacts (*C*_*c*_(*x, x*′)) from the POLYMOD study^22^ and others^23,24^. The final ingredient in *M*_*homo*_, *M*_*rand*_ were the probabilities derived from the vaccine homophily analysis reported above. In the appendix, we show that *M*_*homo*_(*v, x*; *v*^′^, *x*^′^) = [*ϕ*_=_ *C*_=_(*x, x*^′^) + *ϕ*_*c*_ *ωC*_*c*_(*x, x*^′^))*P*(*v*^′^|*v, x, x*^′^) where *ϕ*_=_(resp. *ϕ*_*c*_) is the percentage of household (resp. community) contacts obeying homophily and *M*_*rand*_(*v, x*; *v*^′^, *x*^′^) = ((1 − *ϕ*_=_)*C*_=_(*x, x*^′^) + (1 − *ϕ*_*c*_)*ωC*_*c*_(*x, x*^′^))*P*(*v*^′^|*x*^′^), where *ω* ∈]0.1] is the relative probability of transmission in community contacts relative to household contacts and 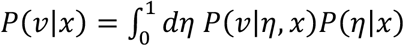 and 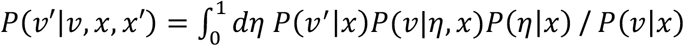. We set *ω* = 0.3 (Ref. ^25^), *ϕ*_=_ = 1(i.e. all household contacts obey homophily) and *ϕ*_*c*_ = 0.25 and investigated alternative choices in the appendix. We assumed leaky vaccine protection reducing susceptibility by a factor E (vaccine efficacy) and testing E=25%, E=50%, E=75%. Finally, to estimate how vaccine homophily impacts transmission, we considered the case where transmission would be purely random, i.e., *ϕ*_=_ = *ϕ*_*c*_ = 0 and computed the corresponding reproduction ratio *R*^(*rand only*)^. The increase in reproduction ratio due to vaccination homophily is then computed as (*R* − *R*^(*rand only*)^)/*R*.

## Results

### Vaccination homophily

The statistical model provided a good fit for the probability of being vaccinated, given age, gender, and reported perceived coverage among friends and family (Fig. 1A and appendix figure S3). We found strong, statistically significant, vaccine homophily in all countries (Fig. 1B,C), as measured by the vaccination odds ratio, i.e., the odds ratio of being vaccinated for an increase in perceived coverage of 10 percentage points. Sweden had the strongest homophily: vaccination O.R. was 1.88 CI=(1.84, 1.91). Hungary had the weakest homophily: vaccination O.R. was 1.44 CI=(1.42, 1.46). The appendix tables 1-3 report the estimated values of the parameters of the statistical model. There was no apparent association between homophily and vaccination coverage (Fig. 1D). In particular, countries in Western and Northern Europe spanned all the observed range of homophily, while all having coverage between 80% and 100%, and most around 90%. Among those countries, however, there was a positive association between homophily and the severity of the first waves of COVID-19, as measured by the total number of COVID-19-attributable deaths in 2020 per 100,000 inhabitants (see appendix figure S4): Pearson correlation with homophily was 0.46 (P=0.047). Notably, the Pearson correlation between homophily and the tightness of restrictions put in place in 2020 (measured by the Stringency Index) was 0.49 (P=0.039) – even stronger than that with surveillance indicators, revealing possible mechanisms linking homophily to the early response against the new pandemic (Fig. 1E). Instead, no correlation existed between mortality and vaccination coverage (Pearson −0.01, P=0.97), or tightness of restrictions and vaccination coverage (Pearson 0.18, P=0.52).

**Fig. 1:**
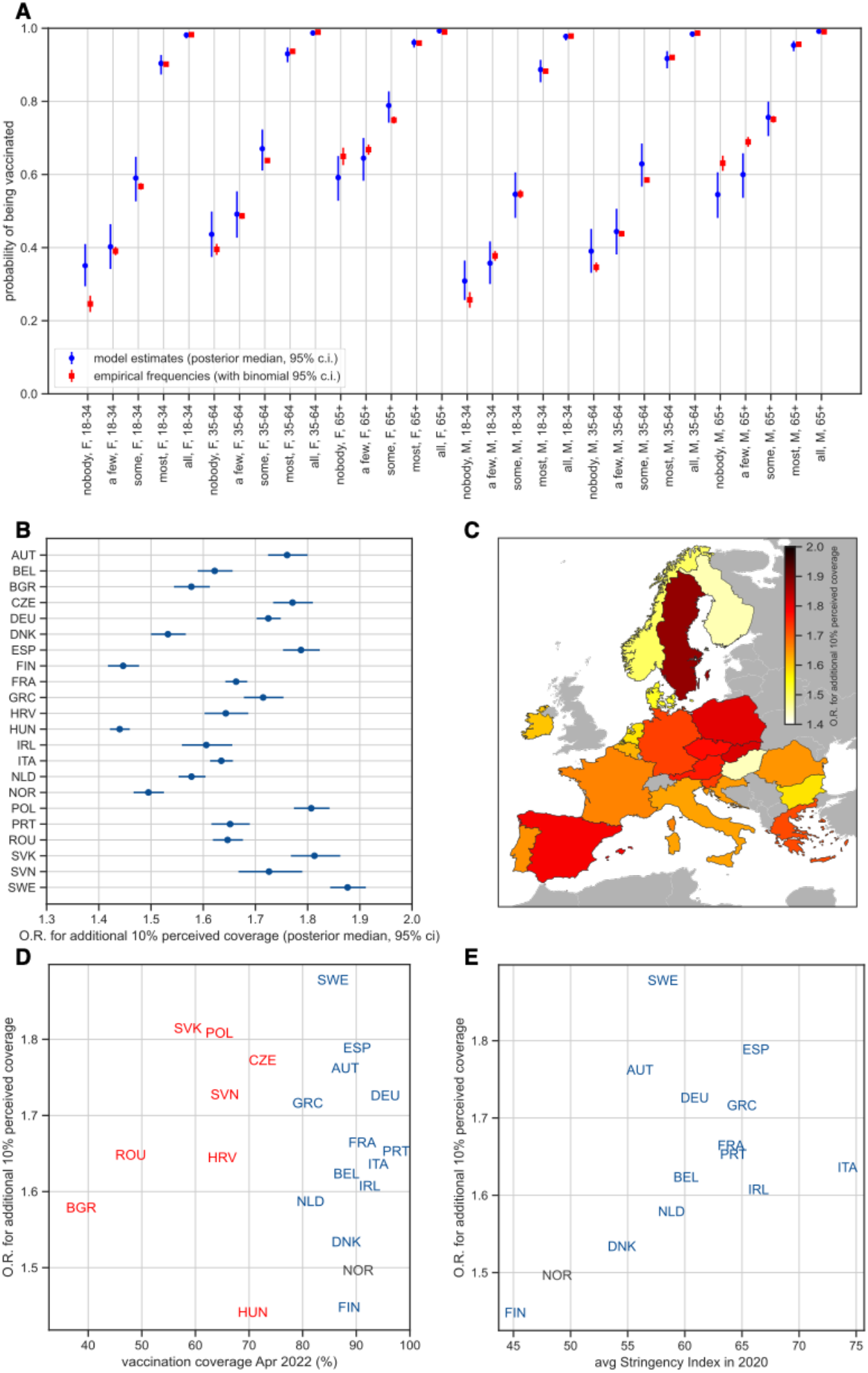
Vaccination homophily. (A) Probability of being vaccinated conditioned on gender, age and reported perceived coverage across the countries under study (see appendix for computation and appendix figure S3 for country-specific probabilities): in blue model estimates reporting posterior median and 95% credibility interval; in red empirical frequencies with binomial 95% confidence intervals. (B) and (C) Posterior estimates of vaccination homophily in terms of posterior vaccination odds ratio for 10% increase in perceived coverage for each country under study: we report median values in B,C and 95% credibility intervals in B. (D) Scatter plot of vaccination coverage (April 2022) vs vaccination homophily. Countries in blue joined the EU before 2004 (Western Europe), those in red in 2004 or after (Eastern Europe). Countries in gray are in the ECDC network but not members of the EU. (E) Scatter plot of Stringency Index (averaged over March-December 2020) and vaccination homophily. Color code is the same as (D).

### Proportion of the reproduction ratio attributable to vaccination homophily

Our model estimated, in each country, the relative difference in the community reproduction ratio of COVID-19 at measured levels of vaccination homophily, and a counterfactual scenario with no homophily, i.e., in which the individual vaccination status is not correlated with the vaccination status of one’s own contacts. Vaccination homophily was responsible for a sizeable fraction of the reproduction ratio in all the countries under study (Fig. 2A). The impact was lowest in Bulgaria (12% of the reproduction ratio), highest in Ireland (18%), at current levels of vaccination coverage, and assuming vaccine to be 50% effective in preventing infection^26^. To estimate this, we used social mixing data from the POLYMOD project^22^, and assumed that vaccination homophily was present among household contacts, and ¼ of community (non-household) contacts, to match the survey question “among your friends and family”. We also explored alternatives to these various assumptions. Alternative social mixing data from Ref.^22–24^ did not significantly change the ranking among countries, and caused the impact of homophily on the reproduction number Rt to increase or decrease by roughly 2 percentage points in each country (appendix figure S5A). Assuming that only household contacts experienced homophily decreased its impact on R_t_ by roughly 3.5 percentage points in each country (appendix figure S5B). Conversely, assuming that ½ of community contacts experienced homophily increased its impact on R_t_ by roughly 3.5 percentage points in each country (appendix figure S5B). The strength of vaccine protection had a strong effect: Very low efficacy (25%) was associated with a low impact of vaccine homophily: 4.5%-to-6.5% of R_t_. High efficacy (75%) raised that to between 22% and 42%. Also, at high vaccine efficacy, the impact on R_t_ increased with vaccine coverage (Fig. 2B).

**Fig. 2:**
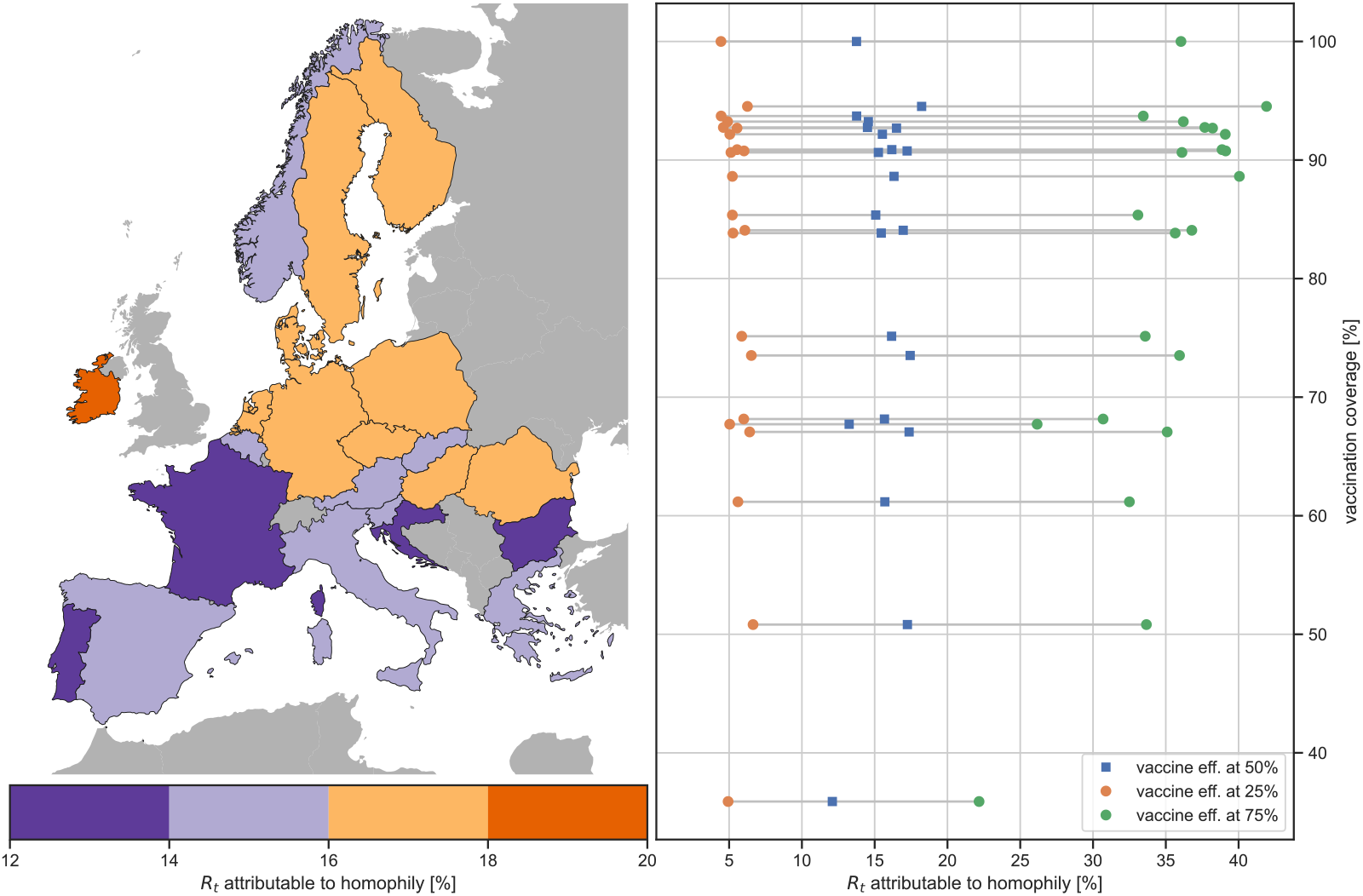
Impact of vaccination homophily on the reproduction ratio. (A) Percentage of the reproduction ratio attributable to vaccination homophily, assuming that vaccination is 50% effective in protecting from infection, contact matrices from the POLYMOD study and that homophily applies to ¼ of non-household contacts. (B) Percentage of the reproduction ratio attributable to vaccination homophily at varying vaccination efficacy.

## Discussion

Vaccination homophily, whereby the contacts of those unvaccinated are themselves likely unvaccinated, characterized all 22 European countries in the first four months of 2022, when primary vaccination campaigns had likely reached the “vaccine compliant” fraction of the population^27^. Despite the high overall vaccination uptake, homophily was strong and unveiled the presence of clusters of unvaccinated people. Vaccination homophily was present in all European countries albeit with different strength. Sweden was the country where homophily was the highest. There, young men reporting that none of their friends were vaccinated had a probability of being vaccinated as low as 20%; those who instead reported that all of those around them were vaccinated had a probability of being themselves vaccinated close to 100% (appendix figure S3). While these differences decreased with less homophily, they were still substantial: in Hungary, where homophily was the lowest, the corresponding probabilities were 40% vs 90%. Vaccination coverage and vaccination homophily had no simple correlation. For instance, vaccination coverage was almost the same in Finland and Sweden, 91% vs 89%, but homophily values were on the opposite side of the spectrum (Fig. 1C). The early evolution of the COVID-19 pandemic may help to explain these differences: early experience with the epidemic could have influenced the willingness to get vaccinated later, although it is not clear in which direction. On the one hand, high incidence and mortality in 2020 could have increased the perception of the risk related to COVID-19, making people more willing to get vaccinated. On the other hand, the exposure to high death and hospitalization counts and the experience of strict restrictions on movement and activities were linked psychological distress^28^, which in turn might have prompted vaccine hesitancy^28,29^. The result of these, and possibly other, competing drives is that, in western Europe, neither early epidemic severity, nor tightness of restrictions, seemed to impact the final vaccination coverage. But interestingly, that was not the case with homophily: a high early burden of COVID-19, and tight restrictions in 2020 were both associated with higher homophily. A possible interpretation of this is that the aforementioned psychological and societal stress contributed to polarizing trust in governments and institutions, which is in turn associated with vaccine acceptance^30^.

Vaccine homophily is a threat to the effectiveness of vaccination campaigns, because clusters of unvaccinated people facilitate viral re-emergence and its circulation in the community, as described for other diseases^9–14^. Here, we showed that vaccination homophily also drives the reproduction ratio (R) of the COVID-19 epidemic up, making control less easy. Our findings indicate that a sizeable fraction of the current reproduction ratio could be attributed to vaccination homophily, ranging from 10% to 20% of the reproduction ratio, assuming that vaccines offer a 50% protection against SARS-CoV-2 acquisition. Differences in homophily, and in age- and gender-stratified vaccination uptake, explained the different impact homophily had on R across countries. Importantly, this means that vaccination uptake alone fails to describe the level of protection in the population – even if stratified on geography, gender and age. Notably, the effect of homophily could be large enough to keep an epidemic wave above the epidemic threshold: if, for instance, homophily is responsible for 20% of the reproduction ratio, and the reproduction ratio is no larger than 1.25, then acting on homophily could stop the epidemic wave. The existence of homophily, and its effect on community circulation, is thus an obstacle to keeping the epidemic under control, but also hints at what public health action should aim at. Indeed, the effectiveness of past and current vaccination campaigns is indisputable: they have been saving lives^31^ and relieving the pressure on the healthcare system without the need of closures and movement restrictions. But the burden of COVID-19 is still high: reported COVID-19 mortality in the period January-October 2022 in the countries under study ranged from 11 to 119 deaths per 100,000 people^1^ and excess mortality was around 10% compared with 2016-2019^32^. Plus, to date, there is no evidence that morbidity and mortality will go down on their own with time as COVID-19 transitions to endemicity^6^. On top of this, post-acute sequelae of COVID-19 may add to its long-term burden^6–8^, if incidence of infections remains high. Current vaccination strategies have little chance to invert this trend and will not keep COVID-19 circulation consistently below the epidemic threshold (R<1), as they focus solely on boosting those already vaccinated: primary vaccinations have stalled for months, and measures to encourage them have been scaled back. Our results instead show that prolonged efforts to increase primary vaccinations, in combination with booster campaigns, would greatly pay off. New primary vaccinations would break the clusters of unvaccinated individuals and reduce homophily, and that would bring down the reproduction ratio, possibly below the epidemic threshold, as our model shows. In addition, we also estimate that the fraction of R attributable to homophily increases if vaccines become more effective, and especially so where uptake is already high. This means that the more effective booster campaigns are, by reaching high coverage and vaccine efficacy^33^, the more it will make sense to concurrently target primary vaccinations, to slash the residual fraction of reproduction ratio and make waves smaller and shorter, or possibly avoid them altogether.

Our study has limitations. It relies on survey respondents accurately reporting their demographic profile, vaccination status and perceived coverage, which may be biased with respect to the actual coverage among one’s own friends and family. The analyzed sample of survey respondents was however large and heterogeneous, decreasing the impact of a perception bias in specific geographic or sociodemographic population strata. Analogously, estimates of the reproduction ratio rely on age- and gender-stratified mixing matrices which themselves rely on survey data. Testing different matrices however gave similar results (appendix figure S5). The estimate of the impact of homophily on R also relied on correctly matching the two data and deciding which social contacts obey the observed homophily: mixing matrices are reported by location (household, community,…), while reported coverage is “among one’s own friends and family”. We tested various assumptions and found little impact on results (appendix figure S5). Another factor impacting the fraction of R explained by homophily was vaccine efficacy in preventing acquisition of SARS-CoV-2. We assumed that protection was the same for all those vaccinated: in reality, it changes with the number of doses, time from last dose and viral variant^26^. We believe, however, that adding this feature to our model would not change its qualitative behavior, and make it over-reliant on data and estimates which are often poorly available. We instead decided to explore a wide range of overall vaccine efficacy to study the impact of booster campaigns (resulting in an increase in average efficacy) on the role of homophily. Our study excludes those younger than 18 years old, not covered by the survey. Their vaccine uptake, however, is much lower than those of adults - 33% vs 86%^1^ -, limiting the bias they might induce when estimating the effect of homophily on COVID-19 circulation. Plus, if this bias does exist, it is likely to underestimate the effect of homophily^34^. Finally, our study suggests that increasing the uptake of primary vaccinations may substantially decrease the circulation and burden of COVID-19, by reducing homophily, but it does not quantify the relationship between new vaccinations and drops in homophily. However, it is reasonable to assume that many new primary vaccinations must occur within cliques of previously unvaccinated individuals, given that most of the population has already been vaccinated, and those vaccinated are surrounded mostly by vaccinated individuals. And vaccinating inside cliques of unvaccinated individuals is what decreases homophily^16,35^.

In Europe, COVID-19 is shifting from an emergent threat to an endemic disease. Vaccination campaigns are keeping the pressure on the healthcare system in check, but the burden of COVID-19 is still considerable, in terms of severe disease, deaths and post-COVID-19 condition. Broadly, the current strategy is to repeatedly boost those who already got vaccinated, giving up on persuading those who have so far refused to. Vaccination campaigns should instead target primary vaccinations too, as our study shows that this has the potential to substantially decrease the circulation of COVID-19 in communities, and its burden. It will not be easy, but the public health benefit may warrant the effort.

## Data Availability

Access to the survey data can be requested for research purposes at https://dataforgood.facebook.com/dfg/docs/covid-19-trends-and-impact-survey-request-for-data-access. The mixing matrices of household and non-household contacts are accessible at http://www.socialcontactdata.org/socrates/. Vaccination coverage data are accessible through the website of the European Centre for Disease Prevention and Control: https://www.ecdc.europa.eu/en/publications-data/data-covid-19-vaccination-eu-eea. The COVID-19 Government Response Tracker is accessible at https://www.bsg.ox.ac.uk/research/research-projects/covid-19-government-response-tracker

## Acknowledgements

We acknowledge Shweta Bansal, Vittoria Colizza and Chiara Poletto for useful discussions. This research is based on survey results from University of Maryland.

## References

1. European Centre for Disease Prevention and Control - Data. European Centre for Disease Prevention and Control https://www.ecdc.europa.eu/en/data.

2. Telenti, A. et al. After the pandemic: perspectives on the future trajectory of COVID-19. Nature 596, 495–504 (2021).

3. Hammond, J. et al. Oral Nirmatrelvir for High-Risk, Nonhospitalized Adults with Covid-19. N. Engl. J. Med. 386, 1397–1408 (2022).

4. Cohen, M. S. Early treatment to prevent progression of SARS-CoV-2 infection. Lancet Respir. Med. 10, 930–931 (2022).

5. Andrews, N. et al. Covid-19 Vaccine Effectiveness against the Omicron (B.1.1.529) Variant. N. Engl. J. Med. 386, 1532–1546 (2022).

6. Bowe, B., Xie, Y. & Al-Aly, Z. Acute and postacute sequelae associated with SARS-CoV-2 reinfection. Nat. Med. 28, 2398–2405 (2022).

7. Phillips, S. & Williams, M. A. Confronting Our Next National Health Disaster — Long-Haul Covid. N. Engl. J. Med. 385, 577–579 (2021).

8. Ballering, A. V. Zon, S. K. R. van Hartman, T. C. olde & Rosmalen, J. G. M. Persistence of somatic symptoms after COVID-19 in the Netherlands: an observational cohort study. The Lancet 400, 452–461 (2022).

9. Edge, R., Keegan, T., Isba, R. & Diggle, P. Observational study to assess the effects of social networks on the seasonal influenza vaccine uptake by early career doctors. BMJ Open 9, e026997 (2019).

10. van den Hof, S., Conyn–van Spaendonck, M. A. E. & van Steenbergen, J. E. Measles Epidemic in The Netherlands, 1999–2000. J. Infect. Dis. 186, 1483–1486 (2002).

11. Parker, A. A. et al. Implications of a 2005 Measles Outbreak in Indiana for Sustained Elimination of Measles in the United States. N. Engl. J. Med. 355, 447–455 (2006).

12. Phadke, V. K., Bednarczyk, R. A., Salmon, D. A. & Omer, S. B. Association Between Vaccine Refusal and Vaccine-Preventable Diseases in the United States: A Review of Measles and Pertussis. JAMA 315, 1149–1158 (2016).

13. Zucker, J. R. et al. Consequences of Undervaccination — Measles Outbreak, New York City, 2018–2019. N. Engl. J. Med. 382, 1009–1017 (2020).

14. Aloe, C., Kulldorff, M. & Bloom, B. R. Geospatial analysis of nonmedical vaccine exemptions and pertussis outbreaks in the United States. Proc. Natl. Acad. Sci. 114, 7101–7105 (2017).

15. Mbah, M. L. N. et al. The Impact of Imitation on Vaccination Behavior in Social Contact Networks. PLOS Comput. Biol. 8, e1002469 (2012).

16. Glasser, J. W., Feng, Z., Omer, S. B., Smith, P. J. & Rodewald, L. E. The effect of heterogeneity in uptake of the measles, mumps, and rubella vaccine on the potential for outbreaks of measles: a modelling study. Lancet Infect. Dis. 16, 599–605 (2016).

17. Burgio, G., Steinegger, B. & Arenas, A. Homophily impacts the success of vaccine roll-outs. Commun. Phys. 5, 1–7 (2022).

18. Gromis, A. & Liu, K.-Y. Spatial Clustering of Vaccine Exemptions on the Risk of a Measles Outbreak. Pediatrics 149, e2021050971 (2021).

19. Global COVID-19 Trends and Impact Survey, in partnership with Facebook | JPSM | Joint Program in Survey Methodology | University of Maryland. https://jpsm.umd.edu/research/global-covid-19-trends-and-impact-survey%2C-partnership-facebook.

20. COVID-19 Government Response Tracker. https://www.bsg.ox.ac.uk/research/research-projects/covid-19-government-response-tracker.

21. Diekmann, O., Heesterbeek, J. a. P. & Roberts, M. G. The construction of next-generation matrices for compartmental epidemic models. J. R. Soc. Interface 7, 873–885 (2010).

22. Mossong, J. et al. Social Contacts and Mixing Patterns Relevant to the Spread of Infectious Diseases. PLOS Med. 5, e74 (2008).

23. Béraud, G. et al. The French Connection: The First Large Population-Based Contact Survey in France Relevant for the Spread of Infectious Diseases. PLOS ONE 10, e0133203 (2015).

24. Van Hoang, T. et al. Close contact infection dynamics over time: insights from a second large-scale social contact survey in Flanders, Belgium, in 2010-2011. BMC Infect. Dis. 21, 274 (2021).

25. Faucher, B. et al. Agent-based modelling of reactive vaccination of workplaces and schools against COVID-19. Nat. Commun. 13, 1414 (2022).

26. Ssentongo, P. et al. SARS-CoV-2 vaccine effectiveness against infection, symptomatic and severe COVID-19: a systematic review and meta-analysis. BMC Infect. Dis. 22, 439 (2022).

27. Neumann-Böhme, S. et al. Once we have it, will we use it? A European survey on willingness to be vaccinated against COVID-19. Eur. J. Health Econ. 21, 977–982 (2020).

28. Aknin, L. B. et al. Policy stringency and mental health during the COVID-19 pandemic: a longitudinal analysis of data from 15 countries. Lancet Public Health 7, e417–e426 (2022).

29. Murphy, J. et al. Psychological characteristics associated with COVID-19 vaccine hesitancy and resistance in Ireland and the United Kingdom. Nat. Commun. 12, 29 (2021).

30. Bollyky, T. J. et al. Pandemic preparedness and COVID-19: an exploratory analysis of infection and fatality rates, and contextual factors associated with preparedness in 177 countries, from Jan 1, 2020, to Sept 30, 2021. The Lancet 399, 1489–1512 (2022).

31. Watson, O. J. et al. Global impact of the first year of COVID-19 vaccination: a mathematical modelling study. Lancet Infect. Dis. 0, (2022).

32. Excess mortality - statistics. https://ec.europa.eu/eurostat/statistics-explained/index.php?title=Excess_mortality_-_statistics.

33. Khoury, D. S. et al. Predicting the efficacy of variant-modified COVID-19 vaccine boosters. 2022.08.25.22279237 Preprint at https://doi.org/10.1101/2022.08.25.22279237 (2022).

34. Nguyen, K. H., Nguyen, K., Mansfield, K., Allen, J. D. & Corlin, L. Child and adolescent COVID-19 vaccination status and reasons for non-vaccination by parental vaccination status. Public Health 209, 82–89 (2022).

35. Alvarez-Zuzek, L. G., Zipfel, C. M. & Bansal, S. Spatial clustering in vaccination hesitancy: The role of social influence and social selection. PLOS Comput. Biol. 18, e1010437 (2022).

