## appendix for "The importance of increasing primary vaccinations against COVID-19 in Europe"

#### Contents

1 Statistical model - vaccination status from perceived coverage 2

2 Reproduction ratio attributable to homophily 4

3 Supplementary Figures and Tables 8

### 1 Statistical model - vaccination status from perceived coverage

For clarity, the model described here is on one country only. The multilevel structure to account for different countries is already described in the main paper. We recap and extend the notation of the main paper:

- $v \in \{0, 1\}$: is vaccinated (binary);
- $j \in \{1, 2, \cdots, K\}$: ordinal predictors of perceived coverage: *who is vaccinated among your friends and family?*. $K$ is the number of ordinal options. Here, $K = 5$, corresponding to the responses: none, a few, some, many, all;
- $x \in \mathbb{R}^M$: variable encoding the additional predictors (age, gender);
- $M \in \mathbb{N}$: number of additional predictors. Here $M = 3$, one for gender, two for age (we used 3 age classes);
- $\eta \in [0, 1]$: perceived coverage.

**Likelihood at given coverage**

$P(v|\eta, x)$: likelihood of being vaccinated given perceived coverage $\eta$, and additional predictors $x$. We assume a classical Bernoulli likelihood with logistic link function (logistic regression):

$$P(v|\eta, x) \sim \text{Bernoulli}\,(p) = vp + (1 - v)(1 - p), \tag{1}$$

$$\log \frac{p}{1 - p} = \alpha + \beta\eta + \gamma \cdot x, \tag{2}$$

with parameters:

- $\alpha$: offset;
- $\beta$: slope of coverage;
- $\gamma \in \mathbb{R}^M$: slopes of the additional predictors;

**Likelihood of having coverage $\eta$, given response**

We define $K-1$ cutpoints $c_m$, with $m \in \{1, 2, \cdots, K-1\}$, and, formally, $c_0 = 0$ and $c_K = 1$. Then we assume that response $j$ corresponds to coverage in the interval $[c_{j-1}, c_j)$, and coverage is uniformly distributed inside that interval:

$$P(\eta|j, x) = \frac{1}{c_j - c_{j-1}} \theta(\eta - c_{j-1}) \theta(c_j - \eta), \tag{3}$$

where $\theta$ is the usual Heaviside theta.

**Final likelihood**

We combine the two previously defined likelihoods as follows:

$$P(v|j, x) = \int_0^1 \mathrm{d}\eta \, P(v|\eta, x) P(\eta|j, x). \tag{4}$$

This gives

$$P(v|j, x) = 1 - v + (2v - 1) \frac{1}{\beta(c_j - c_{j-1})} \log\left(\frac{1 + e^{\alpha + \gamma \cdot x + c_j \beta}}{1 + e^{\alpha + \gamma \cdot x + c_{j-1} \beta}}\right), \tag{5}$$

which can also be rewritten as

$$P(v = 1|j, x) = \frac{1}{\beta(c_j - c_{j-1})} \log\left\{1 + \mathcal{E}(\alpha + \gamma \cdot x + \beta c_{j-1}) \left[e^{\beta(c_j - c_{j-1})} - 1\right]\right\}, \tag{6}$$

$$P(v = 0|j, x) = 1 - P(v = 1|j, x), \tag{7}$$

where $\mathcal{E}(t) = e^t/(1 + e^t)$ (sigmoid function). We note that $P(v|j, x)$ is finite and well-behaved everywhere:

$$P(v = 1|j, x) \overset{\beta \to 0}{\longrightarrow} \mathcal{E}(\alpha + \gamma \cdot x + \beta c_{j-1}); \tag{8}$$

$$P(v = 1|j, x) \overset{c_{j-1} \to c_j}{\longrightarrow} \mathcal{E}(\alpha + \gamma \cdot x + \beta c_{j-1}). \tag{9}$$

**Parametrization of the cutpoints**

The 2 categories $j = 1$ and $j = K$ correspond to *none* or *all* of the friends/family vaccinated. These are really point-mass probabilities ideally corresponding to $\eta = 0, \eta = 1$. In our framework, we model these by a small interval of width $\epsilon$, so that $\eta = 0$ is modelled as $\eta < \epsilon$ and $\eta = 1$ as $\eta > 1 - \epsilon$. $\epsilon$ is given a narrow prior (see after). We parametrize the other cutpoints $c_1, c_{K-1}$ using $\epsilon$: $c_1 = \epsilon$ and $c_{K-1} = 1 - \epsilon$. The remaining $K - 2$ cutpoints are parametrized using a $K - 2$-simplex rescaled to sum to $1 - 2\epsilon$. This gives $K - 2$ degrees of freedom for the cutpoints ($K - 3$ from the simplex, plus $\epsilon$).

##### Priors

We used weakly informative priors except for $\epsilon$. The priors of the meta-parameters $\mu_A, \mu_B$ (main paper) were normal distribution with mean $0$, standard deviation $15$. The priors of the meta-parameters $\sigma_A, \sigma_B$ tuning the dispersion of the country-specific values around $\alpha, \beta$ were exponential distributions with rate $5 \cdot 10^{-3}$. The slopes of the demographic characteristics had also normal priors with mean $0$, standard deviation $15$. Finally, for the cutpoints, for $\epsilon$ we used a an exponential prior with rate $20$ truncated in $[0, 1]$, and for the $K-2$-simplex we used a weak prior: a Dirichlet distribution with $1/(K-2)$ in each entry of its concentration vector.

##### MCMC sampling

We ran the MCMC sampler with $6$ chains, each with $10,000$ iterations. We discarded an initial warm-up time of $2,000$ iterations. Figure S2 shows the value of the log-likelihood for each chain, each iteration, indicating convergence.

#### 2 Reproduction ratio attributable to homophily

Let us consider a Susceptible-Infected-Recovered compartmental model structure on age and gender (synthetically identified by the demographic variable $x$ as before), and vaccination status ($v = \{0, 1\}$). The model shall have an overall rate of transmission $\lambda$ and rate of removal $\gamma$. We used mixing matrices from the POLYMOD study [1], Ref. [2] and Ref. [3], and specifically the matrix of household contacts – $C_H(x, x')$ –, and of community contacts – $C_C(x, x')$. Also, we assumed that a fraction $\phi_H$ of household contacts obey vaccine homophily, and likewise a fraction $\phi_C$ of community contacts. The latter are also less transmissible than the former, by a factor $\omega \leq 1$. Finally, the vaccine is assumed to be leaky and reduce susceptibility by a factor $E$ with no effect on transmission. The equations for the number of infected $I(v, x)$ with vaccine status $v$ and demographic characteristics $x$ are thus

$$\frac{dI(v,x)}{dt} = -\gamma I(v,x) + \lambda(1 - Ev)\frac{S(v,x)}{N(v,x)} \sum_{x',v'} [M_{homo}(v,x;v',x') + M_{rand}(v,x;v',x')]\, I(v',x'), \quad (10)$$

where we used the following definitions, splitting the mixing factor that obeys homophily ($M_{homo}$) and that which does not ($M_{rand}$).

$$M_{homo}(v,x;v',x') = \left[\phi_H C_H(x,x') + \omega\phi_C C_C(x,x')\right] P(v|v',x,x'); \tag{11}$$

$$M_{rand}(v,x;v',x') = \left[(1-\phi_H) C_H(x,x') + \omega(1-\phi_C) C_C(x,x')\right] P(v|x,x'). \tag{12}$$

According to the established theory [4], the next-generation matrix of Eq. (10) is

$$\mathcal{M}(v,x;v',x') = \frac{\lambda}{\gamma}(1-Ev)\left[M_{homo}(v,x;v',x') + M_{rand}(v,x;v',x')\right]. \tag{13}$$

In the counterfactual scenario where transmission is purely random, we set $\phi_H = \phi_C = 0$ in Eq. (11) and Eq. (12) and get

$$\mathcal{M}^{(rand\,only)}(v,x;v',x') = \frac{\lambda}{\gamma}(1-Ev)\left[C_H(x,x') + C_C(x,x')\right] P(v|x,x'). \tag{14}$$

Finally, the largest eigenvalue $R$ of $\mathcal{M}$ gives the reproduction ratio [4], and the largest eigenvalue $R^{(rand\,only)}$ of $\mathcal{M}^{(rand\,only)}$ gives the reproduction ratio when transmission is purely random. The fraction of the reproduction ratio attributable to homophily is thus $(R - R^{(rand\,only)})/R$, from which it is clear that $\lambda, \gamma$ cancel out and need not be estimated – compare Eq. (13) and Eq. (14).

Now, the last step is to compute the conditional probabilities in Eq. (11) and Eq. (12). We will proceed in steps.

**Step I:** $P(v|x,x')$

We define the function $g_1$ of $v, x$:

$$g_1(v,x) = \int_0^1 \mathrm{d}\eta\, P(v|\eta,x)P(\eta|x). \tag{15}$$

Making the assumption that $v$ does not depend on $x$: $P(v|x,x') = P(v|x)$, and decomposing $P(v|x)$ through $\eta$, we find that

$$P(v|x,x') = g_1(v,x). \tag{16}$$

**Step II:** $P(v|v',x,x')$

We manipulate $P(v'|v,x,x')$ by decomposing through $\eta$ and using Bayes' theorem:

$$P(v|v',x,x') = \int_0^1 \mathrm{d}\eta\, P(v|\eta,x,x')P(\eta|v',x,x') \tag{17}$$

$$= \int_0^1 \mathrm{d}\eta\, P(v|\eta,x)P(v'|\eta,x,x')\frac{P(\eta|x,x')}{P(v'|x,x')} \tag{18}$$

$$= \frac{1}{g_1(v',x')}\int_0^1 \mathrm{d}\eta\, \eta P(v|\eta,x)P(\eta|x). \tag{19}$$

Defining

$$g_2(v,x) = \int_0^1 \mathrm{d}\eta\, \eta P(v|\eta,x)P(\eta|x), \tag{20}$$

We finally get

$$P(v|v',x,x') = \frac{g_2(v,x)}{g_1(v',x')}. \tag{21}$$

Eq. 16 and Eq. 21 mean that we computing the required probabilities now requires computing $g_1, g_2$.

**Step III:** $P(\eta|x)$

Our model assumes that $\eta$ is uniformly distributed within categorical bins. So

$$P(\eta|x) = \sum_j \frac{q_j(x)}{c_j - c_{j-1}} \theta(\eta - c_{j-1})\theta(c_j - \eta). \tag{22}$$

The terms $q_j(x)$ are estimated from the data, using the weights provided [5], and are the probability that a person with characteristics $x$ has perceived coverage $j$.

**Step IV: computing** $g_1, g_2$

First, we note that $P(v = 1|\eta, x)$ – the probability of being vaccinated conditioned on $x, \eta$ – comes straight from the definition of the statistical model in Eq. (2):

$$\log \frac{p(\eta,x)}{1-p(\eta,x)} = \alpha + \beta\eta + \gamma \cdot x, \tag{23}$$

where for brevity we wrote $p(\eta, x) = P(v = 1|\eta, x)$. The probability of being vaccinated is Bernoulli–distributed (Eq. (1)), so $P(v = 0|\eta, x) = 1 - p(\eta, x)$.

We now define, and compute, the following two integrals:

$$m_j^{(1)}(x) = \int_{c_{j-1}}^{c_j} \mathrm{d}\eta\, p(\eta,x) \tag{24}$$

$$= \frac{\log\left[1 + e^{\alpha+\beta c_j+\gamma\cdot x}\right] - \log\left[1 + e^{\alpha+\beta c_{j-1}+\gamma\cdot x}\right]}{\beta} \tag{25}$$

$$m_j^{(2)}(x) = \int_{c_{j-1}}^{c_j} \mathrm{d}\eta\, \eta p(\eta,x) \tag{26}$$

$$= \frac{c_j \log\left[1 + e^{\alpha+\beta c_j+\gamma\cdot x}\right] - c_{j-1}\log\left[1 + e^{\alpha+\beta c_{j-1}+\gamma\cdot x}\right]}{\beta} + \tag{27}$$

$$+ \frac{\mathrm{Li}_2\left(-e^{\alpha+\beta c_j+\gamma\cdot x}\right) - \mathrm{Li}_2\left(-e^{\alpha+\beta c_{j-1}+\gamma\cdot x}\right)}{\beta^2},$$

where $\mathrm{Li}_2$ is the classical dilogarithm (or Spence's function), defined, for example, as $\mathrm{Li}_2(z) = -\int_0^z \mathrm{d}t\, \log(1-t)/t$. These integrals will become useful in the following.

We compute $g_1(v, x)$:

$$g_1(v,x) = \sum_j \frac{q_j}{c_j - c_{j-1}} \int_{c_{j-1}}^{c_j} \mathrm{d}\eta \; [1 - v + (2v-1)p(\eta, x)]$$
$$= 1 - v + (2v-1) \sum_j \frac{q_j}{c_j - c_{j-1}} m_j^{(1)}(x). \tag{28}$$

We then compute $g_2(v, x)$:

$$g_2(v,x) = (1-v)\bar{\eta} + (2v-1) \sum_j \frac{q_j}{c_j - c_{j-1}} m_j^{(2)}(x), \tag{29}$$

where $\bar{\eta}$ is simply the mean value of the perceived coverage from Eq. (22).

This completes the computation of the next-generation matrices: Eq. (28), Eq. (29) go into Eq. (16), Eq. (21), and those in turn go into Eq. (11), Eq. (12).

### 3 Supplementary Figures and Tables

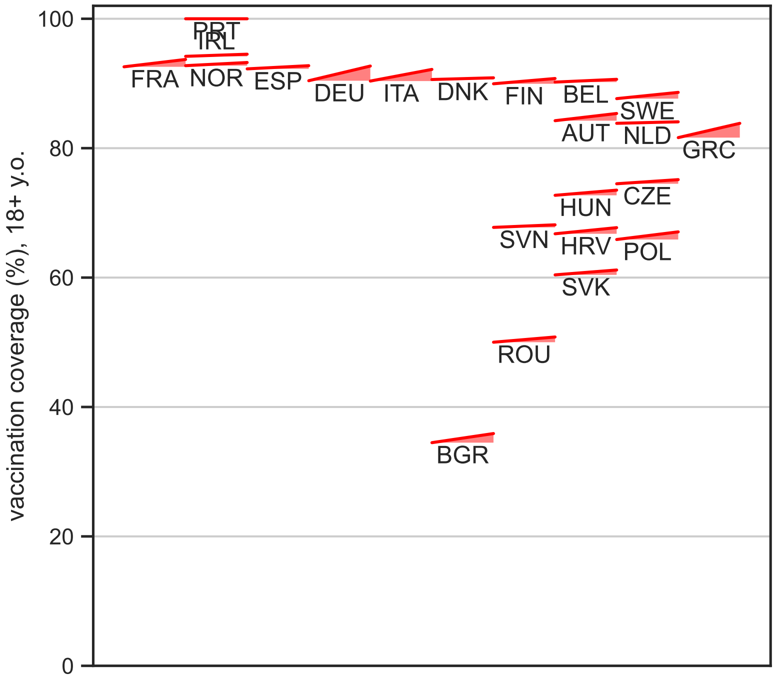


Figure S1: **Country vaccination coverage during the study period**. Each segment represent a country. The left end of the segment is vaccination coverage at the start of the study period (Jan 1, 2022). The right end of the segment is the vaccination coverage at the end of the study period (Apr 30, 2022). Vaccination coverage is the proportion of those aged 18 years old or more having received at least one dose.

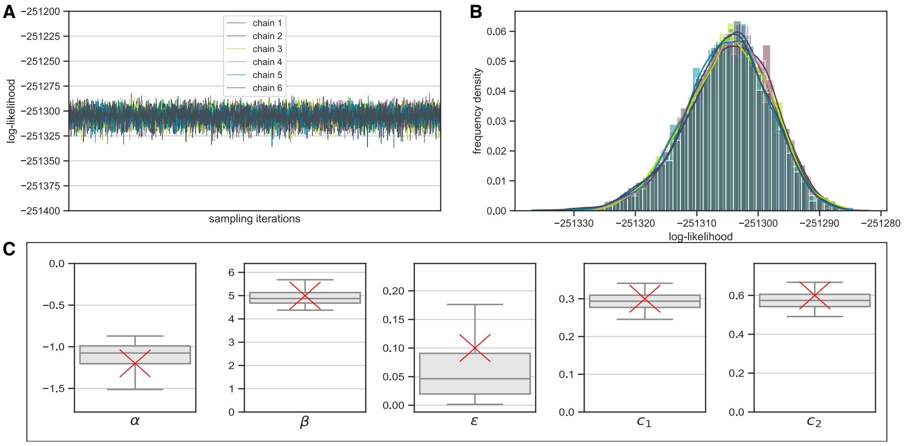


Figure S2: **Model diagnostics**. A) shows the log-likelihood of each MCMC chain, along the sampling iterations (after warm-up). B) shows the histogram of log-likelihood values in each chain. C) Synthetic data (1-country) are generated using the parameter values depicted in red. The model is then fitted to this data, providing the parameter estimates shown as gray boxplots (median, box for 1st and 3rd quartile, whiskers for 95% credibility interval).

| parameter | value |
|---|---|
| $\mu_A$ | -1.0325 (-1.2947, -0.7749) |
| $\sigma_A$ | 0.5823 (0.4341, 0.8308) |
| $\mu_B$ | 5.0439 (4.6866, 5.3990) |
| $\sigma_B$ | 0.7612 (0.5703, 1.0950) |
| $\gamma_1$ | 0.1926 (0.1756, 0.2058) |
| $\gamma_2$ | 0.3636 (0.3431, 0.3796) |
| $\gamma_3$ | 0.9944 (0.9651, 1.0171) |
| $c_1$ | 0.0883 (0.0772, 0.0992) |
| $c_2$ | 0.0886 (0.0774, 0.0994) |
| $c_3$ | 0.3970 (0.3886, 0.4052) |
| $c_4$ | 0.9117 (0.9008, 0.9228) |

Table S1: **Estimated parameters (I)**. Posterior median and $95\%$ credibility interval of the parameters of the statistical model to estimate vaccination homophily.

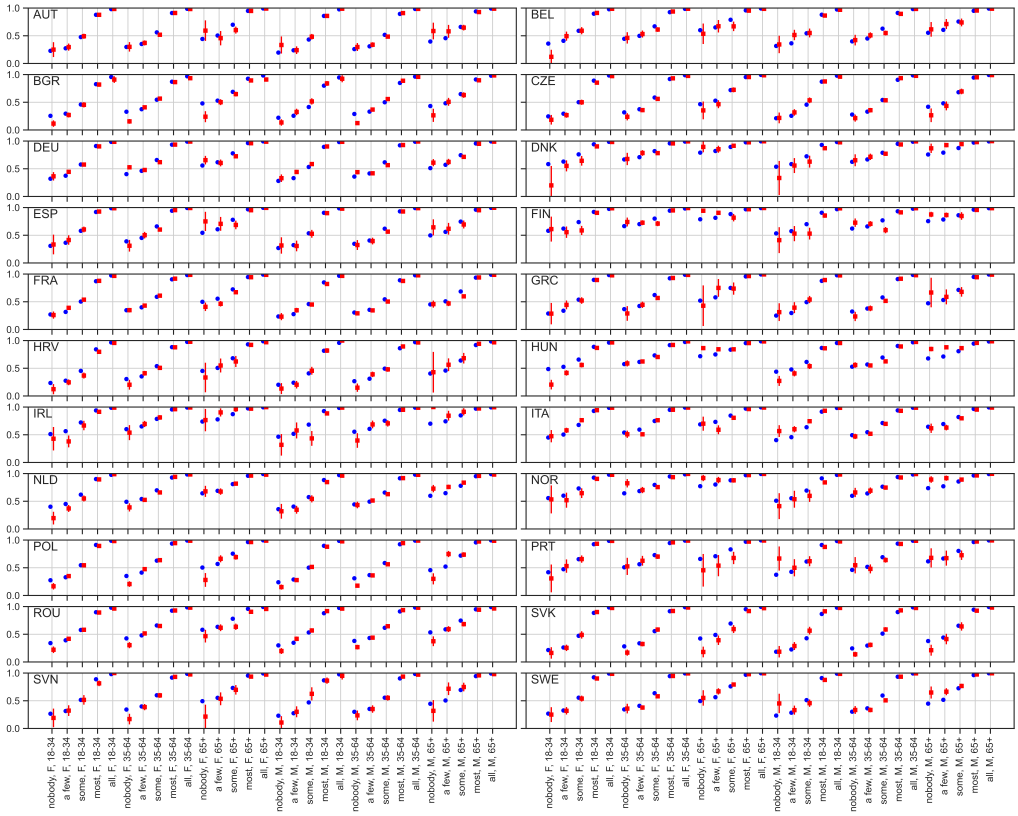


Figure S3: **Probability of being vaccinated**. Probability of being vaccinated conditioned on gender, age and reported perceived coverage, for each country under study. Figure 1A in the main paper reports average probabilities across countries. Model estimates reporting posterior median and 95% credibility interval in blue; Empirical frequencies with binomial 95% confidence intervals in red.

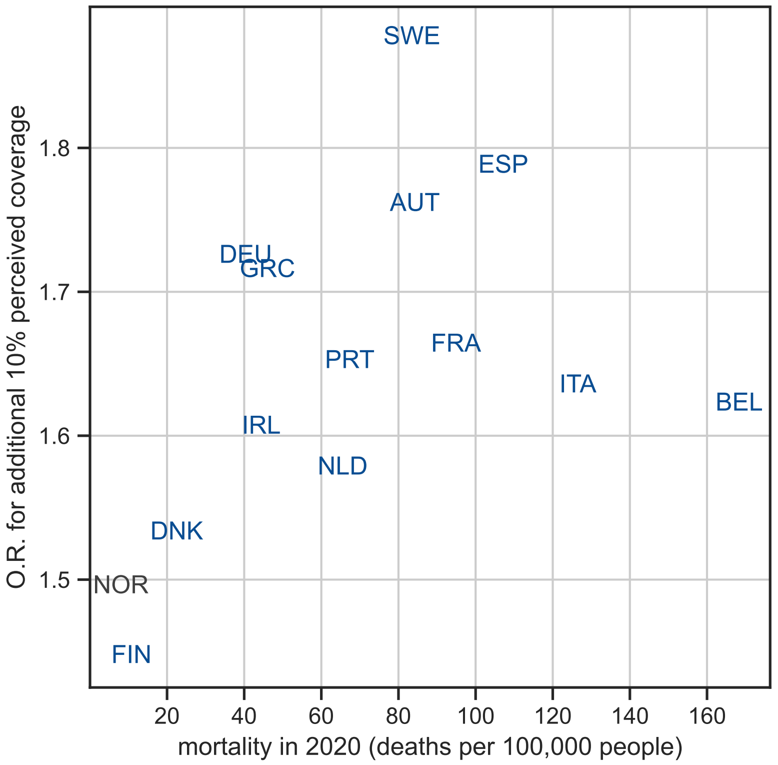


Figure S4: **COVID-19 mortality in 2020 vs homophily**. For homophily, the posterior median odds ratio for 10% increase in perceived coverage is reported. Countries in blue joined the EU before 2004 (Western Europe), those in red in 2004 or after (Eastern Europe). Countries in gray are in the ECDC network but not members of the EU.

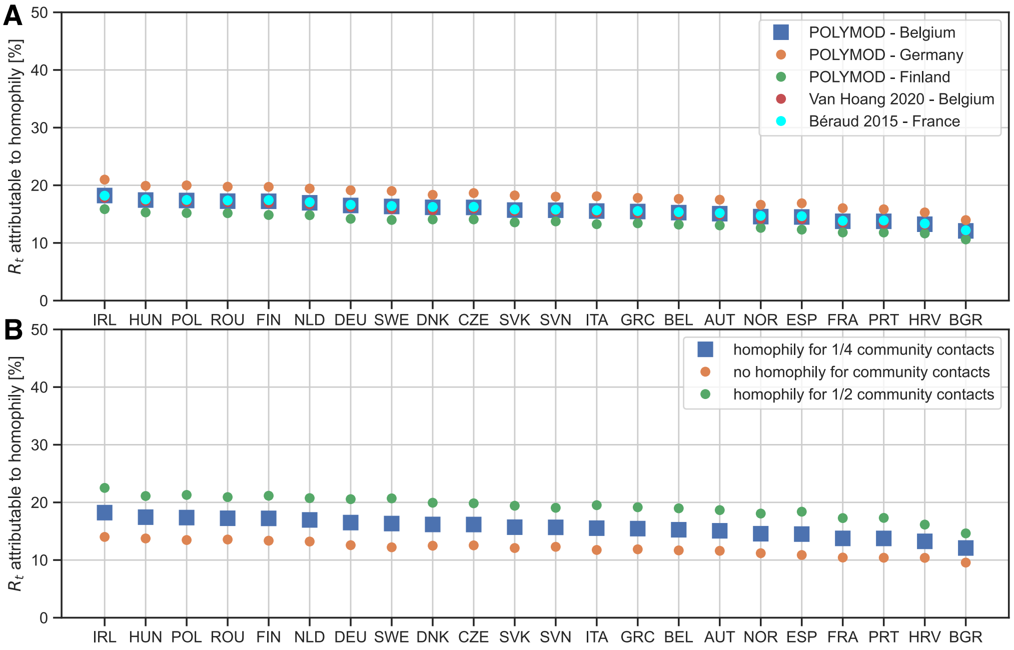


Figure S5: **Impact of homophily on $R_t$: sensitivity**. A) Estimated fraction of $R_t$ attributable to homophily, when changing the data source of the mixing matrices. References: POLYMOD [1], Béraud [3], Van Hoang [2]. B) Estimated fraction of $R_t$ attributable to homophily, when changing the proportion of community (non-household) contacts that obey homophily (parameter $\phi$). In both panel, the blue squares indicate the data and values used in the main paper.

| parameter | value |
|---|---|
| $\alpha$ AUT | -1.6656 (-1.7765, -1.5590) |
| $\alpha$ BEL | -0.9875 (-1.1010, -0.8784) |
| $\alpha$ BGR | -1.4677 (-1.5557, -1.3827) |
| $\alpha$ CZE | -1.5738 (-1.6780, -1.4703) |
| $\alpha$ DEU | -1.1863 (-1.2647, -1.1100) |
| $\alpha$ DNK | -0.0392 (-0.1566, 0.0783) |
| $\alpha$ ESP | -1.2586 (-1.3722, -1.1447) |
| $\alpha$ FIN | -0.0282 (-0.1314, 0.0767) |
| $\alpha$ FRA | -1.4106 (-1.4869, -1.3368) |
| $\alpha$ GRC | -1.3384 (-1.4594, -1.2236) |
| $\alpha$ HRV | -1.5969 (-1.7156, -1.4825) |
| $\alpha$ HUN | -0.4097 (-0.4775, -0.3441) |
| $\alpha$ IRL | -0.3508 (-0.4936, -0.2077) |
| $\alpha$ ITA | -0.6103 (-0.6892, -0.5316) |
| $\alpha$ NLD | -0.7923 (-0.8783, -0.7087) |
| $\alpha$ NOR | -0.1379 (-0.2460, -0.0285) |
| $\alpha$ POL | -1.4282 (-1.5218, -1.3379) |
| $\alpha$ PRT | -0.7394 (-0.8709, -0.6113) |
| $\alpha$ ROU | -1.0747 (-1.1562, -0.9932) |
| $\alpha$ SVK | -1.7565 (-1.8769, -1.6438) |
| $\alpha$ SVN | -1.4526 (-1.5961, -1.3168) |
| $\alpha$ SWE | -1.4792 (-1.5839, -1.3783) |

Table S2: **Estimated parameters (II)**. Posterior median and $95\%$ credibility interval of the parameters of the statistical model to estimate vaccination homophily. Continues from Tab. S1.

| parameter | value |
|---|---|
| $\beta$ AUT | 5.6571 (5.4468, 5.8782) |
| $\beta$ BEL | 4.8364 (4.6344, 5.0460) |
| $\beta$ BGR | 4.5572 (4.3435, 4.7785) |
| $\beta$ CZE | 5.7158 (5.5044, 5.9336) |
| $\beta$ DEU | 5.4503 (5.3190, 5.5879) |
| $\beta$ DNK | 4.2684 (4.0538, 4.4893) |
| $\beta$ ESP | 5.8068 (5.6123, 6.0065) |
| $\beta$ FIN | 3.6918 (3.4882, 3.9010) |
| $\beta$ FRA | 5.0858 (4.9614, 5.2144) |
| $\beta$ GRC | 5.3928 (5.1758, 5.6177) |
| $\beta$ HRV | 4.9667 (4.7157, 5.2257) |
| $\beta$ HUN | 3.6462 (3.5153, 3.7796) |
| $\beta$ IRL | 4.7370 (4.4411, 5.0426) |
| $\beta$ ITA | 4.9140 (4.7819, 5.0491) |
| $\beta$ NLD | 4.5588 (4.3998, 4.7275) |
| $\beta$ NOR | 4.0213 (3.8294, 4.2178) |
| $\beta$ POL | 5.9162 (5.7300, 6.1086) |
| $\beta$ PRT | 5.0169 (4.7982, 5.2423) |
| $\beta$ ROU | 4.9872 (4.8144, 5.1645) |
| $\beta$ SVK | 5.9501 (5.6986, 6.2198) |
| $\beta$ SVN | 5.4582 (5.1140, 5.8210) |
| $\beta$ SWE | 6.2944 (6.1151, 6.4793) |

Table S3: **Estimated parameters (III)**. Posterior median and $95\%$ credibility interval of the parameters of the statistical model to estimate vaccination homophily. Continues from Tab. S2.

### References


[1] Joël Mossong et al. “Social Contacts and Mixing Patterns Relevant to the Spread of Infectious Diseases”. en. In: *PLOS Medicine* 5.3 (Mar. 2008). Publisher: Public Library of Science, e74. ISSN: 1549-1676. DOI: 10.1371/journal.pmed.0050074.

[2] Thang Van Hoang et al. “Close contact infection dynamics over time: insights from a second large-scale social contact survey in Flanders, Belgium, in 2010-2011”. In: *BMC Infectious Diseases* 21.1 (Mar. 2021), p. 274. ISSN: 1471-2334. DOI: 10.1186/s12879-021-05949-4.

[3] Guillaume Béraud et al. “The French Connection: The First Large Population-Based Contact Survey in France Relevant for the Spread of Infectious Diseases”. en. In: *PLOS ONE* 10.7 (2015). Publisher: Public Library of Science, e0133203. ISSN: 1932-6203. DOI: 10.1371/journal.pone.0133203.

[4] O. Diekmann, J. a. P. Heesterbeek, and M. G. Roberts. “The construction of next-generation matrices for compartmental epidemic models”. In: *Journal of The Royal Society Interface* 7.47 (June 2010), pp. 873–885. DOI: 10.1098/rsif.2009.0386.

[5] Neta Barkay et al. “Weights and Methodology Brief for the COVID-19 Symptom Survey by University of Maryland and Carnegie Mellon University, in Partnership with Facebook”. In: *arXiv:2009.14675 [cs]* (Oct. 2020). arXiv: 2009.14675.
